# Meta-Analytic Evidence of Depression and Anxiety in Eastern Europe during the COVID-19 Pandemic

**DOI:** 10.1101/2021.06.21.21259227

**Authors:** Stephen X. Zhang, Saylor O. Miller, Wen Xu, Allen Yin, Bryan Z. Chen, Andrew Delios, Rebecca Kechen Dong, Richard Z. Chen, Roger S. McIntyre, Xue Wan, Senhu Wang, Jiyao Chen

## Abstract

**Objective:** To perform a systematic and meta-analysis on the prevalence rates of mental health symptoms including anxiety and depression during the COVID-19 pandemic in the general population in Eastern Europe, as well as three select sub-populations: students, general healthcare workers, and frontline healthcare workers.

**Data sources:** Studies in PubMed, Embase, Web of Science, Psycinfo, and medRxiv up to February 6, 2021.

**Eligibility criteria and data analysis:** Prevalence rates of mental health symptoms in the general population and key sub-populations during the COVID-19 pandemic in Eastern Europe. Data were pooled using a random-effects meta-analysis to estimate the prevalence rates of anxiety and depression.

**Results:** The meta-analysis identifies and includes 21 studies and 26 independent samples in Eastern Europe. Poland (n=4), Serbia (n=4), Russia (n=3), and Croatia (n=3) had the greatest number of studies. To our knowledge, no studies have been conducted in eleven Eastern European countries including Hungary, Slovakia, and Slovenia. The pooled prevalence of anxiety in 18 studies with 22 samples was 30% (95% CI: 24% – 37%) and pooled prevalence of depression in 18 studies with 23 samples was 27% (95% CI: 21% – 34%).

**Implications:** The cumulative evidence from the meta-analysis reveals high prevalence rates of clinically significant symptoms during the COVID-19 pandemic in Eastern Europe. The findings suggest evidence of a potential mental health crisis in Eastern Europe during the ongoing COVID-19 pandemic. Our synthesis also reveals a relative lack of studies in certain Eastern European countries as well as high heterogeneities among the existing studies, calling for more effort to achieve evidence-based mental healthcare in Eastern Europe.

**Highlights:**

- The pooled prevalence of anxiety and depression were 30% and 27% in Eastern Europe, respectively.

**Trial registration:** CRD42020224458

## 1. INTRODUCTION

The COVID-19 pandemic (1), with more than 170 million confirmed cases and 3.5 million deaths by May 2021 (2), has taken a toll on mental health, due to fear of illness and hospitalization, the effects of social isolation, and economic downturn (2, 3). To assess mental health during the COVID-19 pandemic, several meta-analyses have polled the prevalence of mental health symptoms in several countries or regions, such as China (4), Southeast Asia (5), and South Asia (6). These meta-analytical studies have provided crucial evidence on mental health in specific regions and uncovered important heterogeneity to enable evidence-based healthcare in those regions. However, the literature still lacks meta-analytical evidence on the prevalence of mental health symptoms in Eastern Europe – a vast region that has been affected by the COVID-19 pandemic in the past year. Eastern Europe has struggled to manage the pandemic, and has suffered from high mortality and morbidity rates. Mental health research has historically overlooked Eastern Europe (7), where mental health epidemiology is still regarded with intense stigma and direct evidence on the topic remains scarce (8). Even prior to the pandemic, Eastern Europe has been relatively weak in identifying people with mental health symptoms (7). Furthermore, recent changes in healthcare systems and lack of per capita funding for community mental health resources presents some unique issues in mental health practice at the system level in Eastern Europe (7). Such issues have resulted in a lack of evidence-based mental health practices (8).

This study aims to address this knowledge gap by presenting the first meta-analysis to quantify the prevalence of anxiety and depression during the COVID-19 pandemic in Eastern Europe. We performed a systematic review of the prevalence of anxiety and depression of the general population as well as healthcare workers (HCW) during the COVID-19 pandemic in Eastern European countries. Such meta-analytical pooled prevalence of mental health symptoms provides crucial evidence to enable evidence-based healthcare policies and resource deployment and also creates opportunities for decision-making on prevention.

## 2. METHODOLOGY

This systematic review and meta-analysis follows the Preferred Reporting Items for Systematic Reviews and Meta-Analyses (PRISMA) statement 2019 and is registered in the International Prospective Register of Systematic Reviews (PROSPERO: CRD42020224458) (9).

### 2.1 Data sources and database search strategy

A comprehensive literature search was performed in the following databases: Web of Science, PUBMED, EMBASE, and medRxiv based on keywords shown in Appendix 1 with Boolean operators.

### 2.2 Selection criteria

The articles were selected based on the following criteria:

1. Design: Cohort or cross-sectional
2. Context: COVID-19 pandemic
3. Population: Including adult populations from the general population, general students, medical students, frontline HCWs, or general HCWs
4. Outcome: Anxiety, depression, or insomnia
5. Instrument: Validated measurement tools or scales of anxiety, depression, and insomnia including cut-off scores.
6. Language: English
7. Region: This meta-analysis includes the following Eastern European countries based on the EuroVoc definition of Eastern Europe: Albania, Armenia, Azerbaijan, Belarus, Bosnia and Herzegovina, Bulgaria, Croatia, Czech Republic, Georgia, Hungary, Kosovo, Moldova, Montenegro, North Macedonia, Poland, Romania, Russia, Serbia, Slovakia, Slovenia, Turkey and Ukraine.
8. Time: between February 1, 2020 and February 2021

We excluded articles with the following criteria:

1. Measurements: Non-validated mental health instruments or non-validated cut-off scores.
2. Population: Specific adult populations such as COVID-19 patients, inpatients, or adults under quarantine, children, or adolescents.
3. Methodological approaches: Non-primary studies including reviews, meta-analyses, qualitative studies, case studies, interventional studies, interviews, or news reports.

### 2.3 Empirical study selection

Two coders (BZC & AD) independently reviewed titles and abstracts of empirical studies retrieved through initial screening criteria. Conflicts of eligibility were referred to a third coder (RKD). The articles included after the title and abstract screening underwent a full-text evaluation.

### 2.4 Data extraction

A comprehensive screening protocol was developed. The following variables were collected from each study: author, title, country, starting and ending dates of data collection, study design, population, sample size, respondent rate, female proportion rate, age range and mean, outcome, outcome level, instruments, cut-off scores, and prevalence. If possible, we coded the prevalence at the severity of mild above, moderate above, and severe. For those studies that reported the mild, moderate, and severe prevalence, we converted them into mild above, moderate above, and severe prevalence. For these studies that only reported the overall prevalence, we specified their severity if their cut-off points were reported.

The protocol was followed by two independent coders in pairs (WX & AY, BZC & AD, RZC & SM). The corresponding authors of empirical studies without prevalence data or missing essential data were contacted by a designated researcher (WX). The reason(s) for emailing authors and excluding studies were recorded. After both coders in each pair had independently coded their studies, they then cross-checked their information and discussed possible differences. If disagreements remained, a third coder (SM) settled disagreements after independently coding the study.

### 2.5 Bias risk

Risk of bias was assessed at the empirical article collection and meta-analysis level. Throughout the data collection process, two coders used the Mixed Methods Appraisal Tool (MMAT) to independently determine the appropriateness of measurement tools, the risk of non-response bias, and the sample representativeness of each empirical study (10). A quality score ranging from 0 to 7 was assigned to each empirical study. A MMAT quality score higher than 6 indicated low bias risk, a score between 5 and 6 indicated medium risk, and a score below 5 indicated high bias risk (10). The MMAT scores were compared using a standardized cross-check protocol. A final check of inter-coder consistency was performed by a third independent coder (RKD). A sensitivity analysis was conducted to assess the risk of bias of the study.

### 2.6 Methods of analysis

Stata 16.1 was used to pool rates of anxiety and depression, using metaprop (11). The random-effects model was used to calculate pooled estimates of outcome prevalence between populations. The I^2^ statistic was used to calculate variance difference from effect sizes in order to quantify heterogeneity rather than sampling error (12). Subgroup analyses were performed on population groups (i.e., general population, students, general HCWs, and frontline HCWs), mental health disorder (i.e., anxiety, depression, and insomnia), outcome severity (i.e., mild above, moderate above, severe). We also performed subgroup analysis on EU (European Union) membership, i.e. EU countries vs. Non-EU countries. Lastly, we did subgroup analysis by regions, i.e., the greater Balkan region known more formally as Southeastern Europe and the rest. The greater Balkan region of southeastern Europe includes Albania, Bosnia and Herzegovina, Bulgaria, Croatia, Kosovo, Montenegro, North Macedonia, Romania, Serbia, Slovenia, and Turkey. The remaining Eastern European countries, including Czech Republic, Poland, Russia, and Ukraine, were categorized as Non-Southeastern Europe.

## 3. RESULTS

### 3.1 Screening results

A PRISMA flowchart (Figure 1) illustrates the systematic review process, which is part of a large research effort to examine the prevalence of mental health symptoms across regions and countries during the COVID-19 pandemic. A total of 6949 studies were identified in the search. Of these studies, 3603 were duplicates and were excluded. The initial screening of 3346 studies produced 684 studies eligible for further full-text evaluation. Through detailed full-text evaluation, 524 studies were excluded. Two rounds of emails were sent to the authors of studies with missing data for the meta-analyses. Prevalence data from the email responses was added to 8 of the 29 empirical studies, bringing the total number of empirical studies for the meta-analyses to 168. Of the 168 studies, 23 empirical studies covered Eastern Europe (13-35). As there were only two studies that examined the prevalence of insomnia (19, 22), insomnia was excluded from further analysis. The meta-analysis included the remaining 21 studies with 26 unique samples that reported 87 prevalence rates. Some empirical studies include multiple independent samples. For example, Stojanov et al. surveyed frontline HCWs and general HCWs (17).

**Figure 1.**
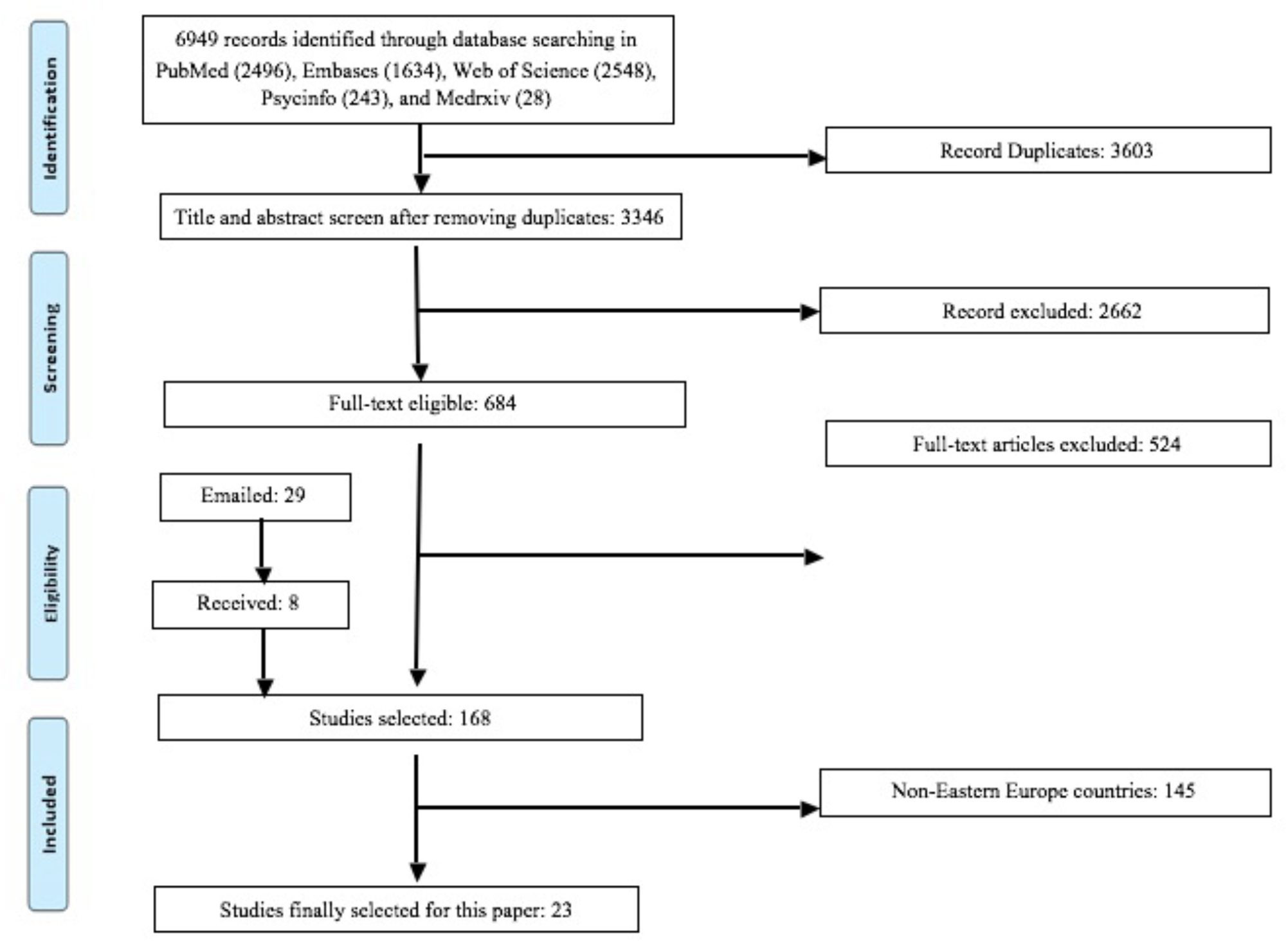
A PRISMA flow diagram. As there were only two studies on insomnia, they were excluded. The final number of studies included in the meta-analysis is 21.

**Figure 2A.**
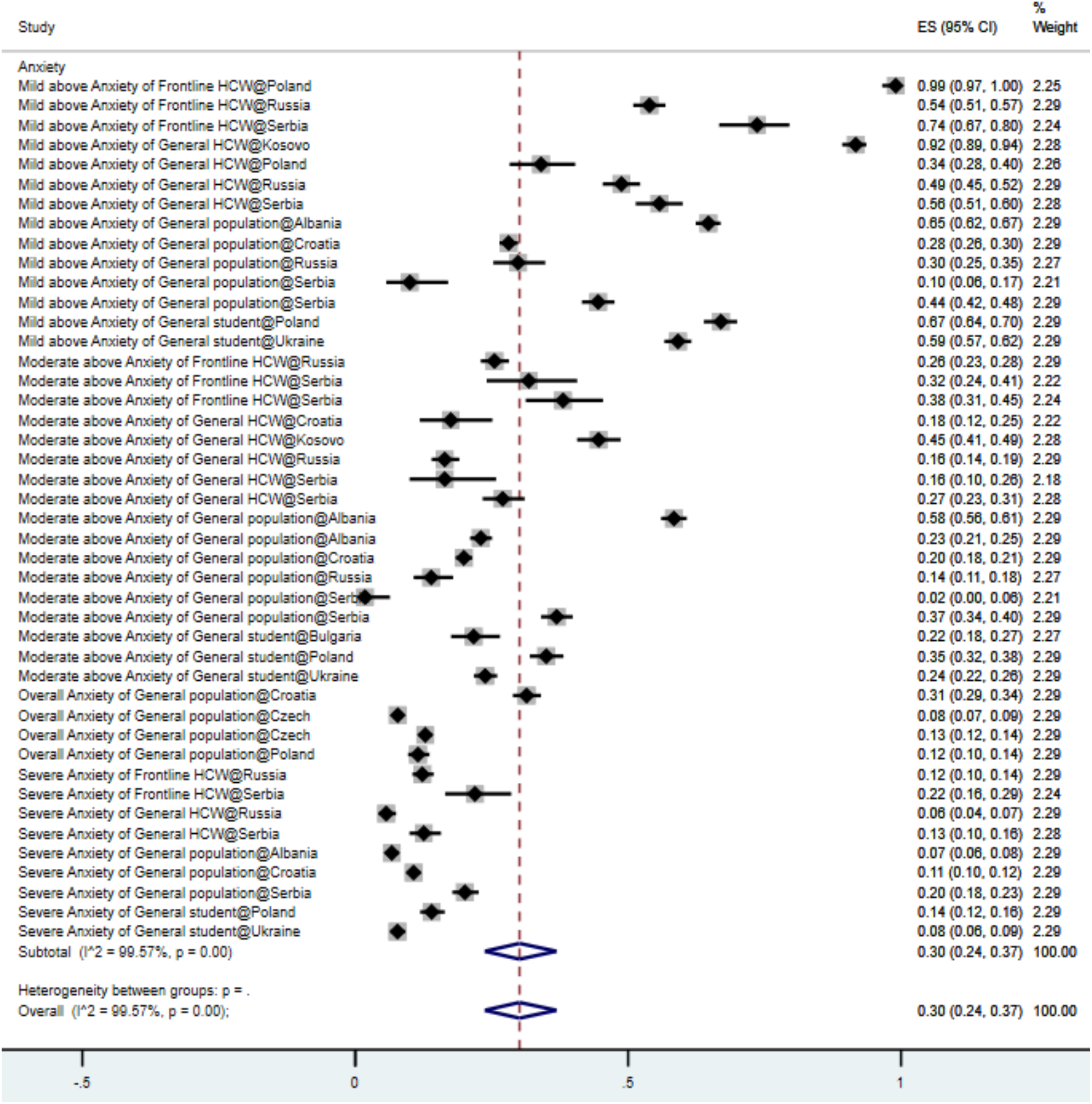
Forest plot of anxiety prevalence. The square markers indicate the prevalence of anxiety at the different levels for different populations. The size of the marker correlates to the inverse variance of the effect estimate and indicates the weight of the study. The diamond data marker indicates the pooled prevalence. The vertical dashed line represents the line of null effect.

**Figure 2B.**
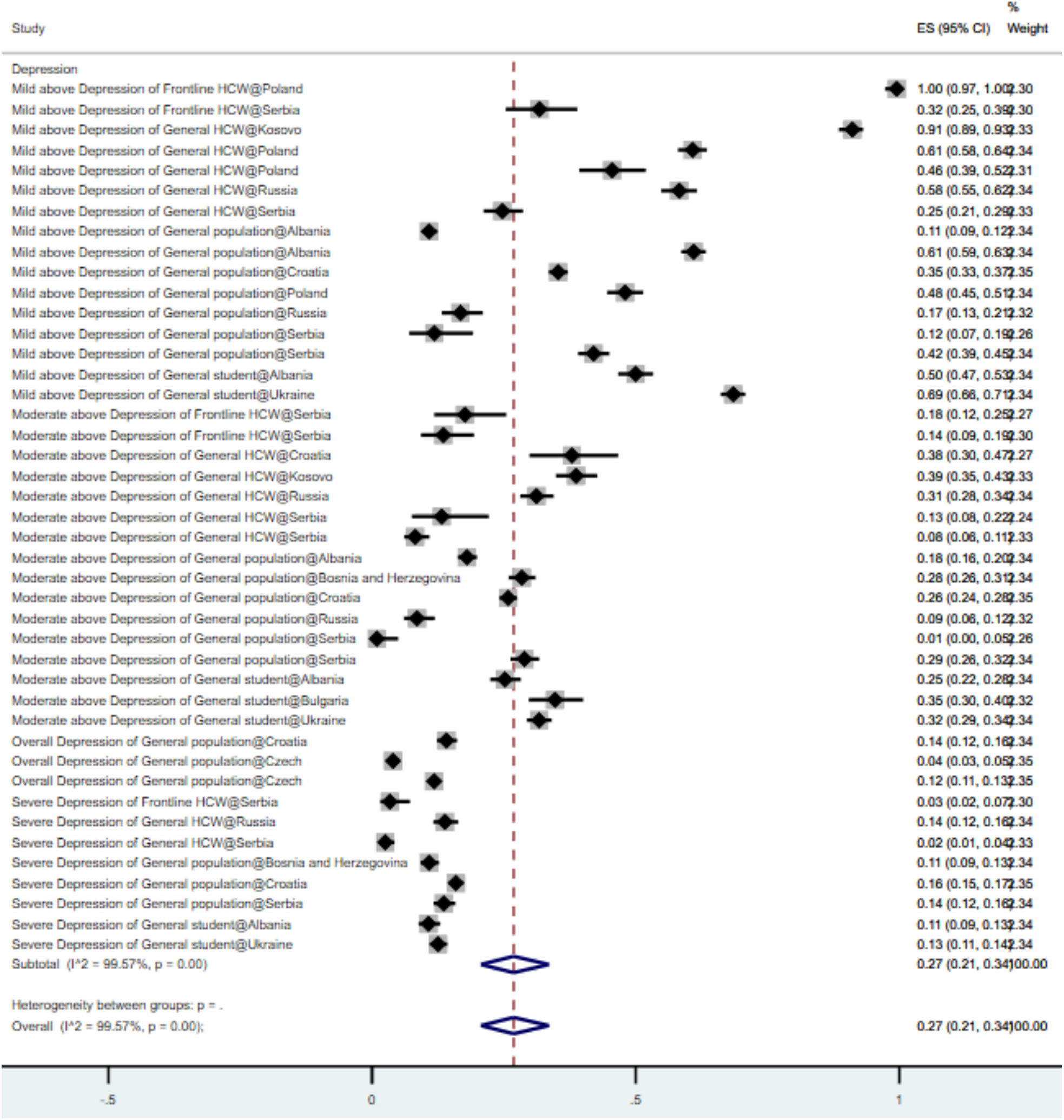
Forest plot of depression prevalence. The square markers indicate the prevalence of depression at the different levels for different populations. The size of the marker correlates to the inverse variance of the effect estimate and indicates the weight of the study. The diamond data marker indicates the pooled prevalence. The vertical dashed line represents the line of null effect.

### 3.2 Characteristics of included studies

Table 1 shows the characteristics of the 21 studies. The countries that had the most studies are Poland and Serbia (19.1%, n=4), followed by Croatia and Russia (14.39%, n=3), Albania (9.5%, n=2), and one study each from Bosnia and Herzegovina, Bulgaria, Czech Republic, Kosovo, and Ukraine. Samples were distributed between populations as follows: general population (42.3%, n=11), general HCWs (26.9%, n=7), general (i.e. non-medical) students (15.4%, n=4), and frontline HCWs (15.4%, n=4). The prevalence of mental health symptoms was found for the following categories: mild above (34.5%, n=30), moderate above (37.9%, n=33), severe above (19.6%, n=17), and overall (8.1%, n=7). Most of the studies (95.7%, n=20) used cross-sectional designs. The median percentage of female respondents was 68.8% with a range of 0% to 100%. The response rates ranged from 0.7% to 98.5% with a median of 56.7%. The sample size ranged from 83 to 3306 with a median of 843 respondents.

**Table 1.** Characteristics of the studies on mental health in Eastern Europe during the COVID-19 pandemic.

| Characteristics | Total number of studies/samples* | Percent | Level of analysis |
| --- | --- | --- | --- |
| *Overall* | 21/26 |  |  |
| *Population* |  |  | Study |
| General population | 9 | 42.86% |  |
| General HCWs | 5 | 23.81% |  |
| General students | 4 | 19.05% |  |
| Frontline HCWs | 3 | 14.28% |  |
| *Outcome#* |  |  | Prevalence |
| Anxiety | 44 | 50.57% |  |
| Depression | 43 | 49.43% |  |
| *Severity#* |  |  | Prevalence |
| Above mild | 30 | 34.48% |  |
| Above moderate | 33 | 37.93% |  |
| Above severe | 17 | 19.54% |  |
| Overall | 7 | 8.05% |  |
| *Sampling country* |  |  | Study |
| Albania | 2 | 9.52% |  |
| Bosnia and Herzegovina | 1 | 4.76% |  |
| Bulgaria | 1 | 4.76% |  |
| Croatia | 3 | 14.29% |  |
| Czech | 1 | 4.76% |  |
| Kosovo | 1 | 4.76% |  |
| Poland | 4 | 19.05% |  |
| Russia | 3 | 14.29% |  |
| Serbia | 4 | 19.05% |  |
| Ukraine | 1 | 4.76% |  |
| *Quality* |  |  | Study |
| High | 3 | 14.29% |  |
| Medium | 18 | 85.71% |  |
| *Study design* |  |  | Study |
| Cohort | 1 | 4.76% |  |
| Cross-sectional | 20 | 95.24% |  |
| *Publication* |  |  | Study |
| Preprint | 2 | 9.52% |  |
| Published | 29 | 90.48% |  |
|  | Mean (Median) | Range |  |
| *Number of participants* | 971 (843) | 83 – 3306 | Sample |
| *Female proportion* | 69.2% (68.8%) | 43.1% – 88.6% | Sample |
| *Response rate* | 54.2% (56.7%) | 0.7% – 98.5% | Sample |

\* A study may include multiple independent samples.

### 3.3 Mental health outcome prevalence

A random-effects meta-analysis model showed the pooled prevalence of depression of 18 empirical studies (13, 15-22, 24-35) (including 23 samples and 43 prevalence rates) was 27% (95% CI: 21% – 34%, I^2^: 99.6%) (Table 2). This pooled prevalence represents a total of 22,195 respondents. Several depression instruments were used: most frequently the Depression, Anxiety, and Stress Scale (DASS-21) (52.2%), followed by Patient Health Questionnaire (PHQ)-9 (30.4%), Beck Depression Inventory (BDI) (4.4%), Hospital Anxiety and Depression Scale (HADS) (4.4%), and Brief Symptom Inventory (BSI) (4.4%).

**Table 2.** The pooled prevalence rates of mental health symptoms by subgroups of population, outcome, severity, region, and quality.

| First-level subgroup | Second-level subgroup | Prevalence (%) | 95%CI (%) | I<sup>2</sup> (%) | P value |
| --- | --- | --- | --- | --- | --- |
| Outcome | Aggregated | 28% | 24–33 | 99.6 | 0.00 |
|  | Anxiety | 30% | 24–37 | 99.6 | 0.00 |
|  | Depression | 27% | 21–34 | 99.6 | 0.00 |
| Population | Frontline HCWs | 41% | 23–60 | 99.4 | 0.00 |
|  | General HCWs | 33% | 22–45 | 99.5 | 0.00 |
|  | General population | 21% | 16–26 | 99.7 | 0.00 |
|  | General students | 31% | 20–44 | 99.6 | 0.00 |
| Severity | Above mild | 52% | 43–60 | 98.5 | 0.00 |
|  | Above moderate | 24% | 20–28 | 98.1 | 0.00 |
|  | Above severe | 11% | 9–13 | 95.2 | 0.00 |
|  | Overall | 12% | 8–18 | 99.1 | 0.00 |
| EU membership | EU countries | 34% | 27–42 | 99.7 | 0.00 |
|  | Non-EU countries | 28% | 22–35 | 99.8 | 0.00 |
| Region | Southeastern Europe | 27% | 22–32 | 99.4 | 0.00 |
|  | Non-Southeastern Europe | 32% | 24–40 | 99.7 | 0.00 |
| Quality | high quality | 28% | 23–32 | 99.8 | 0.00 |
|  | medium quality | 34% | 19–51 | 99.3 | 0.00 |

Note: CI = Confidence Interval. I<sup>2</sup> statistic indicates the heterogeneity across the studies.

The pooled prevalence of anxiety was 30% (95% CI: 24% – 37%, I^2^: 99.6%) (Table 2). Data from 18 studies (13-15, 17-26, 28-35), including 22 samples and 44 prevalence rates), reported anxiety prevalence out of a total of 21,120 participants. The DASS-21 was used most frequently (56.5%), followed by Generalized Anxiety Symptoms 7-items scale (GAD-7) (30.4%), HADS (4.4%), and BSI (4.4%).

The aggregated prevalence of either anxiety or depression in frontline HCWs was 41% (95% CI: 23 – 60%, I^2^: 99.4%) and 33% in general HCWs (95% CI: 22 – 45%, I^2^: 99.4%) (Table 2). The overall prevalence of mental health symptoms in Southeastern Europe countries was lower (27%, 95% CI: 22% – 32%, I^2^: 99.4%) than in non-Southeastern Europe countries (32%, 95% CI: 24% – 40%, I^2^: 99.7%) (Table 2).

Subgroup analysis revealed that while depression prevalence was 34% for both general HCWs (95% CI: 18% – 51%, I^2^: 99.5%) and frontline HCWs (95% CI: 2% – 79%, I^2^: 99.5%), anxiety prevalence was significantly higher among frontline HCWs (46%; 95% CI: 25% – 67%, I^2^: 99.4%) than among general HCWs (33%; 95% CI: 16% – 51%, I^2^: 99.5%) (Table 3). The prevalence of anxiety and depression in student populations was 31% and 32%, respectively. In the general population, prevalence of depression and anxiety was 20% and 22%, respectively (Table 3). European Union (EU) countries in Eastern Europe had a prevalence of 34% (95% CI: 27% – 42%, I^2^: 99.7%), which is a bit higher than that Eastern European countries without EU memberships s (28%; 95% CI: 22% – 35%, I^2^: 99.8%). Southeastern Europe countries (the greater Balkan region) had pooled anxiety and depression prevalence rates of 31% (95% CI: 23% – 40%, I^2^: 99.5%) and 35% (95% CI: 21% – 51%, I^2^: 99.4%), respectively (Table 3). In non-Southeastern Europe countries, prevalence of anxiety and depression was 29% (95% CI: 20% – 40%, I^2^: 99.6%) and 55% (95% CI: 46% – 65%, I^2^: 99.8%) (Table 3).

**Table 3.** Subgroup analyses of anxiety and depression prevalence.

| Groups | Subgroups | Anxiety | Depression |
| --- | --- | --- | --- |
| Number of studies |  | 18 | 18 |
| Number of samples |  | 22 | 23 |
| Number of prevalence rates |  | 44 | 43 |
| Number of participants |  | 21,120 | 22,195 |
| Aggregated |  | **30%**, 95% CI: 24% – 37%,<br>I<sup>2</sup>: 99.6% | **27%**, 95% CI: 21% – 34%,<br>I<sup>2</sup>: 99.6% |
| Population | Frontline HCWs | **46%**, 95% CI: 25% – 67%,<br>I<sup>2</sup>: 99.4% | **34%**, 95% CI: 2% – 79%,<br>I<sup>2</sup>: 99.5% |
|  | General HCWs | **33%**, 95% CI: 16% – 51%,<br>I<sup>2</sup>: 99.5% | **34%**, 95% CI: 18% – 51%,<br>I<sup>2</sup>: 99.5% |
|  | General population | **22%**, 95% CI: 15% – 31%,<br>I<sup>2</sup>: 99.6% | **20%**, 95% CI: 13% – 27%,<br>I<sup>2</sup>: 99.5% |
|  | General student | **31%**, 95% CI: 15% – 50%,<br>I<sup>2</sup>: 99.7% | **32%**, 95% CI: 16% – 50%,<br>I<sup>2</sup>: 99.6% |
| Severity | Above mild | **56%**, 95% CI: 44% – 67%,<br>I<sup>2</sup>: 99.4% | **48%**, 95% CI: 36% – 60%,<br>I<sup>2</sup>: 99.6% |
|  | Above moderate | **26%**, 95% CI: 20% – 32%,<br>I<sup>2</sup>: 98.6% | **22%**, 95% CI: 17% – 26%,<br>I<sup>2</sup>: 96.8% |
|  | Severe | **12%**, 95% CI: 9% – 15%,<br>I<sup>2</sup>: 95.6% | **10%**, 95% CI: 7% – 13%,<br>I<sup>2</sup>: 94.8% |
|  | Overall | **15%**, 95% CI: 8% – 24%,<br>I<sup>2</sup>: 99.2% | **9%**, 95% CI: 4% – 17% |
| EU membership | EU countries | **28%**, 95% CI: 20% – 38%,<br>I<sup>2</sup>: 99.5% | **36%**, 95% CI: 24% – 48%,<br>I<sup>2</sup>: 99.7% |
|  | Non-EU countries | **29%**, 95% CI: 20% – 40%,<br>I<sup>2</sup>: 99.8% | **27%**, 95% CI: 18% – 37%,<br>I<sup>2</sup>: 99.6% |
| Region | Southeastern Europe (Greater Balkan region) | **31%**, 95% CI: 27% – 40%,<br>I<sup>2</sup>: 99.5% | **35%**, 95% CI: 21% – 51%,<br>I<sup>2</sup>: 99.4% |
|  | Non-Southeastern Europe | **29%**, 95% CI: 20% – 40%,<br>I<sup>2</sup>: 99.6% | **55%**, 95% CI: 46% – 65%,<br>I<sup>2</sup>: 99.8% |
| Instruments | GAD-7/PHQ-9 | **34%**, 95% CI: 25% – 44%,<br>I<sup>2</sup>: 99.6% | **35%**, 95% CI: 35% – 57%,<br>I<sup>2</sup>: 99.6% |
|  | DASS-21 | **25%**, 95% CI: 16% – 34%,<br>I<sup>2</sup>: 99.1% | **28%**, 95% CI: 20% – 36%,<br>I<sup>2</sup>: 98.8% |
|  | HADS | **43%**, 95% CI: 17% – 71%,<br>I<sup>2</sup>: 99.1% | **33%**, 95% CI: 7% – 67%,<br>I<sup>2</sup>: 99.7% |
|  | BDI | **5%**, 95% CI: 3% – 9%, I<sup>2</sup>:<br>98.8% | **12%**, 95% CI: 4% – 24%,<br>I<sup>2</sup>: 97.5% |
|  | SDS | NA | **10%**, 95% CI: 3% – 20%,<br>I<sup>2</sup>: 88.8% |

Note: CI = Confidence Interval; I<sup>2</sup> statistic indicates heterogeneity.

### 3.4 Article quality

Study quality was analyzed using the Mixed Methods Appraisal Tool (MMAT). Of the 21 studies, 3 studies (14.29%) were categorized as high quality and 18 studies (85.71%) categorized as medium quality (Table 1). The subgroup analysis suggests the studies with high quality reported lower prevalence of clinically significant symptoms of mental health symptoms in Eastern Europe (Table 2).

### 3.5 Sensitivity analysis

A sensitivity analysis was conducted using a DOI plot and Luis Furuya-Kanamori (LFK) index to detect any publication bias in the meta-analysis. Conventional funnel plots have been previously determined to be inaccurate for meta-analyses of pooled proportion studies (34). Additionally, DOI plots in combination with LFK indices use higher power and sensitivity for bias detection than both funnel plots and Egger’s regression (36). A DOI plot, in addition to a LFK index, can better graphically represent publication bias. An asymmetrical triangle indicates potential publication bias whereas a symmetrical triangle suggests no publication bias (36). A LFK index score within ±1 indicates ‘no asymmetry’. When the LFK index score exceeds ±1 but is within ±2 it indicates ‘minor asymmetry’ and when the score exceeds ±2 ‘major asymmetry’ is indicated. The studies on Eastern Europe, as shown in Figure 3 have ‘minor asymmetry’ based on an index score of 1.50 and therefore minor publication bias is likely. The impact of publication status and sample size was tested and no significant influence was found.

**Figure 3.**
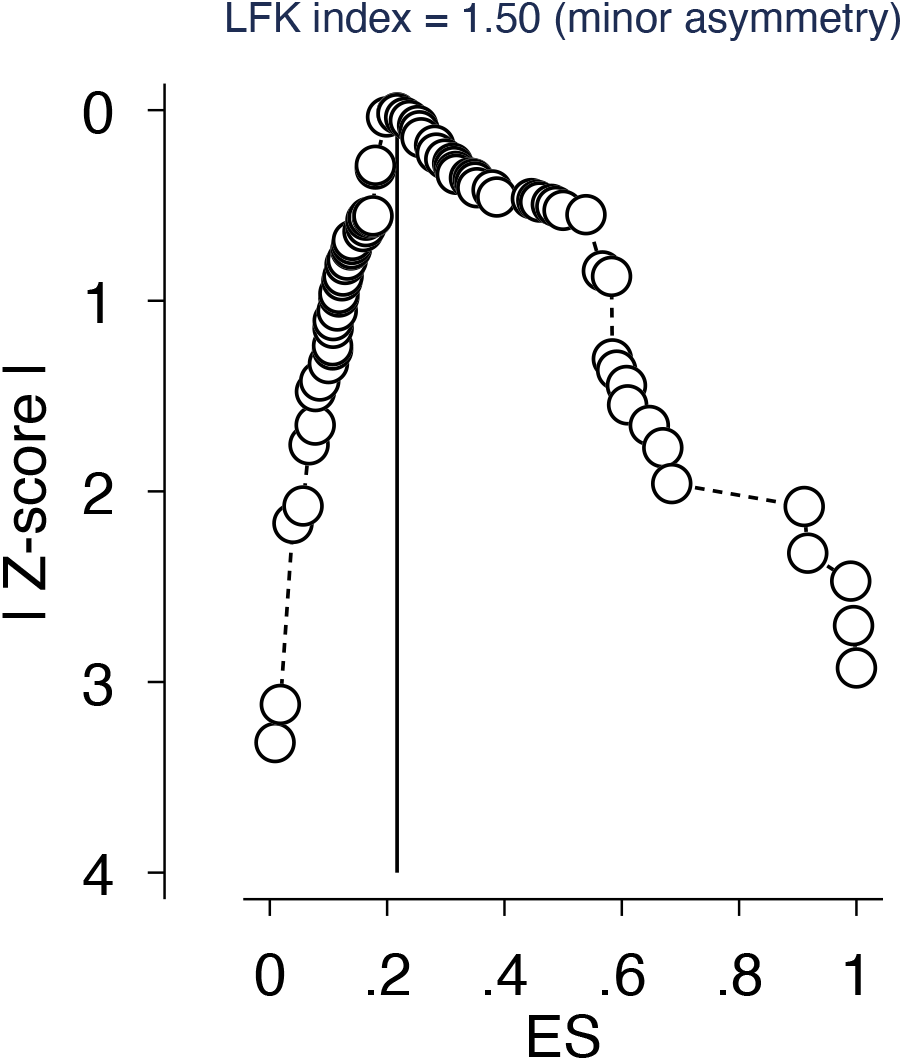
Luis Furuya-Kanamori Index indication of publication bias. Publication bias in the baseline meta-analysis is graphically represented using a DOI plot along with a Luis Furuya–Kanamori (LFK) index score. A score of 1.50 indicates ‘minor asymmetry’ and therefore minor publication bias.

## 4. DISCUSSION

This meta-analysis on 21 empirical studies of 25,246 adults provides the first evidence on the pooled prevalence rates of mental health symptoms in Eastern Europe during the COVID-19 pandemic. The pooled prevalence rates of anxiety and depression in Eastern Europe were 30% and 27% (Table 2). Subgroup analyses revealed several key findings: frontline HCWs indicated higher rates of mental disorder symptoms, especially anxiety (46%, 95% CI: 25% – 67%, I^2^: 99.4%), a high percentage of adults suffered from mild or greater severity of anxiety (56%, 95% CI: 44% – 67%, I^2^: 99.4%), a higher rate of depression was found in non-Southeastern Europe (55%, 95% CI: 46% – 65%, I^2^: 99.8%), and the choice of instruments represents a significant source of heterogeneity on the pooled prevalence of anxiety and depression symptoms.

### 4.1 Comparing with prior meta-analyses

First, we discuss the pooled prevalence rates of this meta-analysis by comparing them with prior meta-analytical findings during the COVID-19 pandemic in other regions as benchmarks. Our pooled prevalence of depression of 27% in Eastern Europe was within the range of similar meta-analyses results in China. Prior meta-analyses of depression in China indicated heterogeneous pooled prevalence ranging from 24% to 32% in adult populations (4, 37-39). Our pooled prevalence of depression in Eastern Europe was significantly lower than prevalence in Southeast Asia (34%, p<0.001) (5), Spain (35%, p <0.001) (40) and Africa (45%, p <0.001) (41) and prevalence rates in a cross-continent meta-analysis including empirical studies from China, India, Iran, Iraq, Italy, Japan, Nepal, Nigeria, Spain, and the UK (34%, p <0.001) (42).

The pooled prevalence of anxiety of 30% in this Eastern Europe meta-analysis was significantly higher than the pooled rates reported in Spain (20%, p <0.001) (40) and was similar to those among the general population in China (30%, p=0.653) (37). However, prevalence of anxiety symptoms was significantly lower in Eastern Europe compared to prevalence in Africa (37%, p <0.001) (40) (41) and Southeast Asia (41%, p <0.001) (5), and to prevalence in a cross-continent meta-analysis (i.e., China, India, Iran, Iraq, Italy, Japan, Nepal, Nigeria, Spain and the UK) (32%, p <0.001) (42).

### 4.2 Subgroup analyses

Our results show that frontline HCWs suffered from anxiety symptoms at a significantly higher rate compared to other populations in Eastern Europe. Overall, frontline HCWs had the highest prevalence of mental health symptoms including anxiety (46%) and depression (34%), followed by general HCWs (anxiety: 33%, depression: 34%) and subsequently students (anxiety: 31%, depression: 32%) and the general population (anxiety: 22%, depression: 20%). This finding indicates heterogeneity of mental health symptoms among distinct populations in Eastern Europe. Comparatively, a meta-analysis in China also found a lower pooled prevalence of anxiety (27%) and depression (20%) among general HCWs compared to prevalence of anxiety (40%) and depression (24%) in frontline HCWs (43). Notwithstanding this finding, it needs to be emphasized that there are heterogeneous results on the prevalence of psychopathology reported in HCWs in China (44).

The prevalence of mental health symptoms during the COVID-19 pandemic is not homogeneous across regions. European Union (EU) countries in Eastern Europe had a prevalence of 34%, which a bit higher than that Eastern European countries without EU memberships at 28%. The greater Balkan region of Southeastern Europe and non-Southeastern Europe exhibited a similar rate of anxiety (31% vs. 29%) but a very different rate of depression symptoms (35% vs. 55%). Such mental health symptom differences provide important evidence for future research directions to offer insight into these significant differences. It is possible that the economic, cultural, social, and political factors of individual countries and broader regions as well as heterogeneous COVID-19 policies, such as the length and stringency of shutdowns, lockdowns, and quarantine, may influence mental health symptoms such as anxiety and depression differentially (45). We also note that the pooled prevalence rates of anxiety and depression are significantly influenced by the choice of the instruments in the primary studies. For example, anxiety prevalence measured by DASS-21 was 25% (95% CI: 16% – 34%, I^2^: 99.1%) and 43% using HADS (CI: 17% – 71%, I^2^: 99.1%), suggesting future research should pay attention to the choice of the instruments.

### 4.3 Implications

The systematic review reveals that eleven Eastern European countries had not been subject to a single study on the topic. Future studies should focus on countries without empirical data including Armenia, Azerbaijan, Belarus, Georgia, Hungary, Moldova, Montenegro, North Macedonia, Romania, Slovakia, and Slovenia. For practical purposes, healthcare organizations in locations without country-level evidence on mental health may use our evidence at the regional level as approximate evidence. These findings also emphasize the importance of further empirical research and subsequent meta-analyses on Eastern Europe countries in order to better prioritize resource allocation.

The understanding of mental disorder prevalence within specific regions can help to create targeted healthcare policy by healthcare organizations such as WHO. The responses to the COVID-19 pandemic have been remarkably homogeneous across governments (45). Available WHO guidance has focused on preventing local progression of infectious diseases rather than achieving regional herd behavior (45). Eastern European mental healthcare is dependent on large psychiatric institutions with an emphasis on in-patient psychiatry (7), which may not be effectively addressing widespread anxiety and depression symptoms. Mental health research has historically been overlooked in Eastern Europe (7), where mental health epidemiology is still regarded with intense stigma and direct evidence on the topic remains scarce (8). Existing stigma, along with a lack of evidence-based community-wide mental health services, may be contributing to high prevalence of mental health disorders. Furthermore, recent changes in healthcare systems and lack of per capita funding for community mental health resources may contribute to the unique situation in Eastern Europe mental health, which still lacks evidence-based mental health practices (7). With this backdrop, the meta-analysis provides quantitative evidence revealing a high prevalence of mental health symptoms in Eastern Europe serve as the basis for inform more targeted healthcare practices, such as evidence-based occupational guidelines which identify and focus on vulnerable populations during acute crises (2).

### 4.4 Study limitations

First, as we only included studies in English, there is an expected language bias. Second, our meta-analysis is limited by the limitations of the empirical studies. Due to the nature of lockdowns and social isolation during the pandemic, many of the studies used convenience samples, reducing the accurate representation of respective populations. Varying tools of data collection used different cut-off scores. In future research, mental health evaluation of a random sample would yield representative data. Additionally, the meta-analysis is limited by the populations and mental health symptoms represented in the available empirical studies. Only two studies covered insomnia, and therefore insomnia was not represented in the meta-analysis. Further studies focused on insomnia prevalence would contribute to a pooled prevalence of insomnia and improve supporting data for evidence-based medical interventions.

### 4.5 Conclusion

Understanding the prevalence of mental health symptoms during the COVID-19 pandemic represents the first step to enable evidence-based medical practices by assessing the mental health situation during the COVID-19 pandemic. We hope the meta-analysis in Eastern Europe can inform mental health practices as well as encourage future research on mental health during the ongoing COVID-19 pandemic.

## Data Availability

The data that support the findings of this study are available from the corresponding author, J.C., upon reasonable request.

## Appendix 1 The search strategy of this systematic review and meta-analysis

Appendix 1: The search strategy of this systematic review and meta-analysis
| Search query | Search topic | Search keywords (titles, abstracts, and subject headings) with Boolean operators |
| --- | --- | --- |
| **1** | Exposure/<br>Context | “Coronavirus” OR “COVID-19” OR “SARS-CoV-2” OR “2019-nCoV” |
| **2** | Outcome of<br>interest | “Depression” OR “Depressive symptoms” OR “Depressive disorder*” OR “Anxiety” OR “Social anxiety” or “Social phobia” OR “Anxiety disorder” OR “Insomnia” OR “Sleep disorder” OR “Depressive disorder*” |
| **3** | Epidemiological<br>phenomenon | “Prevalence” OR “Incidence” OR “rate*” OR “ratio*” OR “Epidemiolog” OR “risk factor” OR “relative risk” OR “odds ratio” OR “risk ratio” OR “disease burden” |
| **4** | Language | English |
| **Final search query** | Intersection of<br>four topics | 1 AND 2 AND 3 AND 4 |

## Author contributions

*All authors declare they meet ICMJE conditions for authorship*. ***SZ, RKD*** *and* ***JC*** *designed the coding guide and method and performed the initial literature search*. ***SZ, SM***, *and* ***JC*** *performed statistical analysis and led the drafting and editing of the article*. **RSM** *edited the article*. **SM, WX, AY, BZC, AD, RKD, RZC** and **XW** *collected the data. All authors approved the final version of the article*.

## Transparency declaration

*The corresponding author affirms this manuscript is an honest, accurate, and transparent account of the study being reported. No important aspects of the study have been omitted and any discrepancies from the study as planned (and, if relevant, registered) have been explained*.

## Ethical approval

*Not applicable*.

## Patient and public involvement

*No patient or public was involved in this systematic review and meta-analysis*.

## Availability of data

*The data that support the findings of this study are available from the corresponding author, J*.*C*., *upon reasonable request*.

